# Effects of Controlled Diets High in and Free of Ultraprocessed Food on the Brain of Emerging Adults

**DOI:** 10.64898/2026.04.30.26352056

**Authors:** Emma H. Leslie, Maria Rego, Monica Ahrens, Wenjing Yu, Mary Elizabeth Baugh, Anastasia Groccia, Rhianna Sullivan, Han Lee, Ryann Kolb, Delbert L. Herald, Valisa E. Hedrick, Kevin P. Davy, Benjamin Katz, Brenda M. Davy, Alexandra G. DiFeliceantonio

**Affiliations:** Graduate Program in Translational Biology, Medicine, and Health, Virginia Tech, Blacksburg, VA, United States; Fralin Biomedical Research Institute, Virginia Tech Carilion, Virginia Tech, Roanoke, VA, United States; Department of Human Nutrition, Foods, and Exercise, Virginia Tech, Blacksburg, VA, United States; Department of Internal Medicine, Division of Medical Informatics, University of Kansas Medical Center, Kansas City, KS; School of Neuroscience, Virginia Tech, Blacksburg, Virginia, USA; Department of Biology, Virginia Tech, Roanoke, Virginia, USA; Department of Human Development and Family Science, Virginia Tech, Blacksburg, Virginia, USA

**Keywords:** /Phrase: ultra processed food (UPF), habitual UPF, functional MRI, young adult

## Abstract

**Objective:** The average American consumes 55% of their daily energy from ultraprocessed foods (UPF) created through industrial processes and additives not used at home. We investigated if a high-UPF diet alters brain response to milkshake compared with a diet free-from UPF (NonUPF) in emerging adults, who are in a critical period for brain development and typically consume high amounts of UPF.

**Methods:** In a randomized controlled crossover trial participants aged 18-25 completed two, 2-week controlled feeding periods including a UPF (81% UPF) and nonUPF (0% UPF) diet. Before and after each diet intervention participants consumed milkshake concomitant with functional magnetic resonance imaging.

**Results:** In the entire cohort, there were no differences between diet conditions in brain response. An exploratory analysis revealed orbitofrontal cortex (OFC) response to milkshake decreased after the UPF diet and increased following the NonUPF diet in adolescents (18-21 years) but not young adults (22-25 years). Habitual UPF intake (gs) was positively associated with OFC response to milkshake independent of diet intervention in all participants.

**Conclusions:** An acute UPF dietary intervention may only alter brain response in adolescents. Further work is needed to determine potential vulnerability of adolescents to changes in dietary UPF on brain response to rewards.

## Introduction

Ultraprocessed foods (UPFs) are combinations of ingredients resulting from a series of industrialized processes that are distinct from their minimally processed counterparts^1–3^. In the United States (US), 55% of daily energy is consumed as UPFs, and UPF intake is substantially higher (∼68% total energy) among emerging adults^1,4–6^. UPF consumption has been linked to adverse health outcomes such as diabetes, cardiovascular disease, weight gain, overweight and obesity, cancer, and all-cause mortality^7–11^. In two randomized controlled feeding trials among adults, participants exposed to an a*d libitum* UPF (81% kcals from UPF) diet consumed more energy compared with an *ad libitum* un/minimally processed (0% kcals from UPF) diet, which led to weight gain^12,13^.

UPF consumption has been linked to poor health outcomes across body systems, including the brain. Pre-clinical studies examining the effects of “junk food”, “cafeteria”, or “western” diets have demonstrated effects on brain function, structure, and receptor distribution^10,14–17^. From these studies, the hippocampus and reward-related circuitry emerge as areas that may be particularly vulnerable to diet-related changes^17–20^. In humans, studies have linked dietary fat, often consumed as a UPF, to changes in cognition^21–23^. Interventions with foods characteristic of UPFs, such as high-fat/high-sugar or sugar sweetened beverages, demonstrate functional brain changes in reward circuitry. ^24,25^. This suggests functional brain changes occur after exposure to constituents common in UPFs, but those studies weren’t designed to explicitly test effects of food processing while holding other dietary factors equal.

Adolescence and young adulthood represent a critical period of life where executive function matures, and independent food choices increase. Importantly, the cortico-limbic system, involved in executive control of eating behavior continues development into the third decade of life^26^. This system, including the hippocampus and prefrontal cortex, is crucial in control of eating behavior^27^. These developing brain areas also appear to be vulnerable to structural and functional effects after dietary intervention^14,15,17^.

Dietary intervention studies in humans are necessary to determine if dietary change during adolescents and young adulthood leads to functional brain changes. To date, no studies have tested the effects of UPF consumption on the brain through a nutrient-matched feeding trial in this age group. Previously, we published findings from the trial described here, demonstrating increased *ad libitum* energy intake after a 2-week 81% UPF diet compared with a 2-week 0% UPF eucaloric diet in adolescents aged 18-21, but not young adults aged 22-25^28^.

Here, we examine the effects of the same trial and cohort on brain response to milkshake via functional magnetic resonance imaging (fMRI). We hypothesized that the blood oxygen level dependent (BOLD) signal would be blunted in reward regions of the brain, such as the striatum and prefrontal cortex, following the UPF diet compared with the NonUPF diet. Further, we hypothesized that blunted response to milkshake after the UPF diet would correlate with increased energy intake at an *ad libitum* buffet meal and eating in the absence of hunger test.

## Methods

Recreationally active participants aged 18-25 completed two 14-day controlled feeding periods in randomly assigned order, with a 4-week washout between diet conditions, as previously described^28,29^. Trial design and conduct was consistent with recommended practices for feeding trials^30^. Diets were eucaloric, matched for macro- and micro-nutrients, and differed in their level of processing (81% UPF, 0% UPF) as classified by NOVA^1–3^. Details on diet development and controlled diet’s energy density, and nutrient contents have been reported^29^. Briefly, diets were comparable to the typical US diet and matched across diet conditions: 50% carbohydrate, 35% fat, 15% protein, and matched in fiber^31^, added sugars, saturated fat, sodium ^31,32^, and overall diet quality (HEI-2015) ^33^. Total energy density was similar (e.g., 2000 kcal menus: UPF, 1.3 kcal/g; non-UPF, 1.2 kcal/g). Non-beverage energy density was 2.1 kcal/100g for the UPF diet and 1.99 kcal/100g for the non-UPF diet^28,29^. Body weight was monitored daily to ensure weight stability. After each diet intervention, participants arrived overnight-fasted the next morning and completed an *ad libitum* breakfast buffet meal followed by eating in the absence of hunger (EAH) snack. Before and after each diet intervention and prior to the *ad libitum* buffet meal, participants completed a milkshake task concomitant with fMRI. The trial design overview is depicted in **Figure 1A**. This trial was registered at ClinicalTrials.gov (NCT05550818) and the Virginia Tech Institutional Review Board (22-253) gave ethical approval for this work.

**Figure 1.**
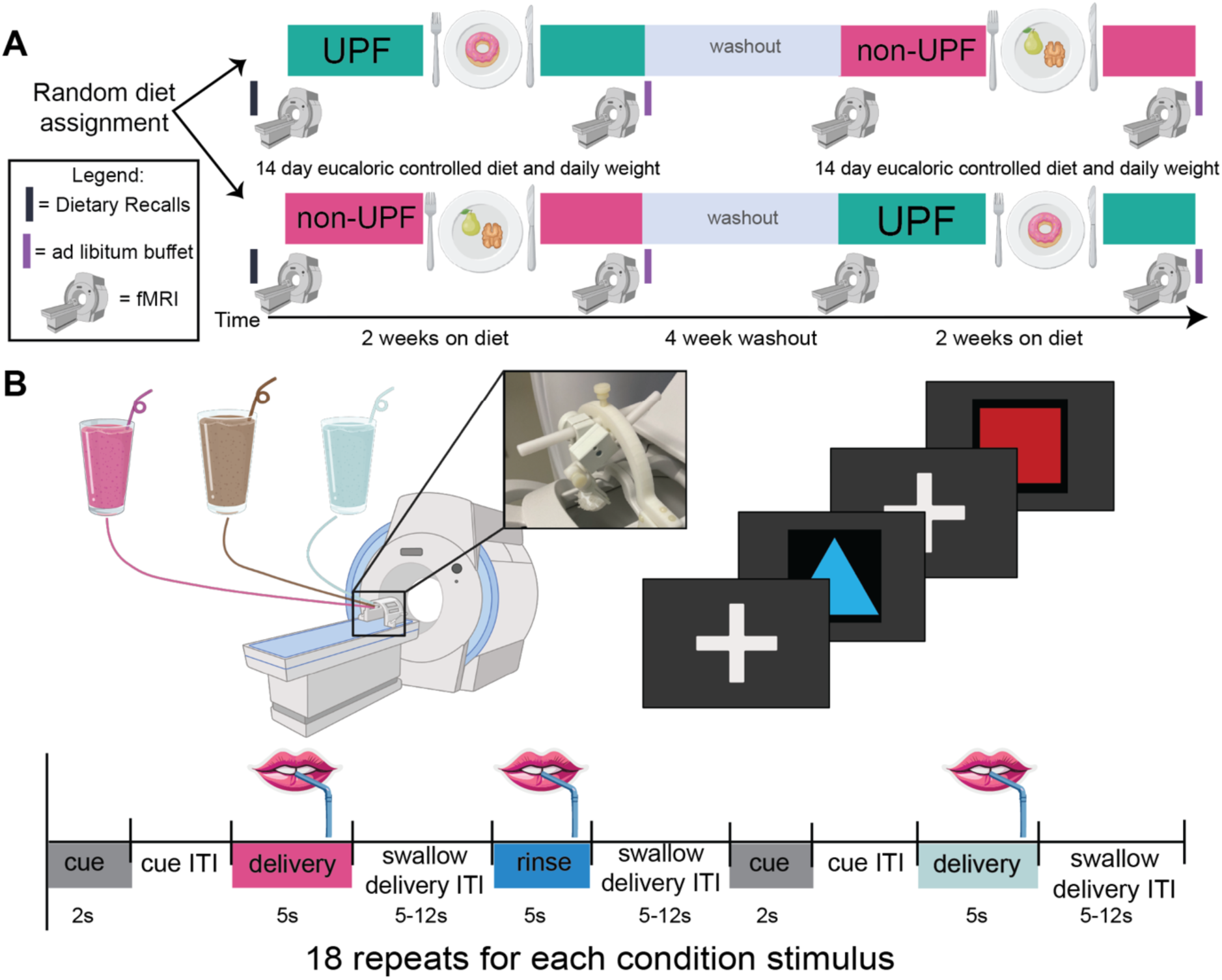
Trial Design. **A.)** Participants complete two 14-day controlled diet periods, in random order, separated by a 4-week washout. The two eucaloric diet conditions were an UPF (81% kcals UPF) and a non-UPF diet (0% kcals UPF). Before and after each diet period, participants complete two runs of a milkshake task with simultaneous fMRI. **B.)** The milkshake is delivered via a pump system and custom manifold that attaches to the head coil. Participants receive a milkshake predictive cue, delivery of milkshake, then a rinse of water interleaved with a tasteless predictive cue and tasteless solution. ITI: inter-trial interval, UPF: ultraprocessed food

## Recruitment and Screening

Recruitment for this trial began February 2023, and data collection was completed in December 2024 (**Supplemental Figure 1**). Participant eligibility was pre-screened using an online survey; potential qualifiers received a trial food list to ensure no allergies or aversions to foods used. Then, an in-person screening was conducted with the following: informed consent procedures; anthropometrics (height, weight, body mass index [BMI]); assessment of dietary cognitive restraint, disinhibition, hunger^34^, physical activity level^29,35^; MRI contraindications, and a 24-hour dietary recall^28,29^. Participants completed a “mock scan” in a simulated MRI environment where they were presented with a shortened version of the task to practice swallowing supine. Two additional dietary recalls were performed via phone, totaling one weekend and two weekdays^29^. Participants that met eligibility and consented to the trial were randomized by 3 stratification factors: age (18-21 or 22-25), sex (male or female), and BMI (18.5-24.9 or 25-29.9 kg/m^2^), determining diet condition sequence according to an *a priori* generated randomization scheme.

## Inclusion Eligibility

In addition to eligibility criteria described in previous manuscripts^28,29^, participants were required to have completed all four fMRI sessions with at least 1 run (out of 2 available runs) within each session passing imaging quality checks. Quality checks included: 1. runs with movement > 2mm in a single direction were excluded and 2. runs where >20% of volumes were flagged as having a change in DVARS (D referring to the temporal derivative, VARS referring to RMS variance over voxels) greater 1.5 standard deviations from the mean were excluded^36^. Out of 28 participants that completed the study, 1 exclusion occurred due to missing data (power outage), 2 exclusions due to equipment malfunction, 1 exclusion due to illness during testing, and 2 exclusions due to quality checks; the final sample presented here is n=22 (**Supplemental Figure 1**).

## Habitual Dietary Intake

Habitual intake was determined using three 24-hour dietary recalls as described previously^28,29^. For 2 participants, 1 recall day was excluded due to lack of detail. Recall data was entered in the Nutrition Data Systems for Research (NDSR 2022, University of Minnesota, Nutrition Coordinating Center). Two trained evaluators, including one RDN (DH), used NDSR output files to score all foods consumed using the NOVA classification system. Discrepancies were addressed and resolved by a third, PhD-level RDN (MEB). Habitual UPF intake in % grams was used to account for habitual intake consistent with published best practices for applying the NOVA system to dietary intake data^3^.

### Ad Libitum Buffet

The morning following each 14-day diet period, participants arrived overnight-fasted to complete an *ad libitum* buffet breakfast meal. Briefly, participants were presented with five items each of UPF and non-UPF items that were comparable in food type (grain, fruit, etc) and were group-level matched on acceptability and texture^28,29^. As previously described, internal state ratings of hunger, fullness, and thirst were completed before and immediately after the *ad libitum* buffet meal. Time spent eating, amount consumed (kcals, grams), eating rate (kcals/min, grams/min), and food selection (UPF, non-UPF) were recorded and are reported elsewhere^28,29^.

## Eating in the Absence of Hunger

When the participant stopped eating the *ad libitum* meal, regardless of if participants used the full allotted time, participants waited a standardized 15 minutes before completing the eating in the absence of hunger (EAH) test that included 3 each of UPF and non-UPF items.

Internal state (hunger, fullness, thirst) was rated immediately before the task, to ensure absence of hunger^28^. The nutritional composition and texture of these items were matched at the condition level (UPF vs non-UPF)^29^. Participants were asked to taste and rate each of the foods and then were asked to eat as much or as little as they would like over the next 15 minutes^28,29^.

## Functional Magnetic Resonance Imaging

Before the diet intervention and prior to the *ad libitum* buffet meal for the post-session, BOLD response to milkshake was acquired via fMRI. All scans were acquired using a Magnetom Prisma whole-body 3T Prisma scanner with 64-channel head coil (Siemens AG, Medical Solutions, Erlangen, Germany). Two, 11-minute echo planar image (EPI) runs were acquired (TR: 1500.0 ms, TE: 34.00 ms, field of view:192 mm, flip angle: 70°, voxel size: 2.0 × 2.0 × 2.0 mm^3^, 72 axial slices, ascending interleaved in-plane acquisition, anterior to posterior phase encoding direction, multi-band acceleration factor 4). Two images with identical parameters but opposite phase encoding (posterior to anterior) were collected to estimate and correct susceptibility-induced distortion. Structural images were collected using a T1-weighted sequence (TR: 2300 ms, TE: 2.32 ms, field of view: 240 mm, flip angle: 8 deg., 0.9 × 0.9 × 0.9 mm3, and 192 sagittal slices).

## Milkshake Task

As previously described, participants underwent two, 11-minute runs of the task where participants receive two stimuli (milkshake vs tasteless), each predicted by a distinct cue^29^. To avoid habituation, participants selected two flavors of milkshake from chocolate, strawberry, and caramel^24,37,38^, made with whole milk and commercially available syrup (Hershey’s). Stimuli were delivered via pump system through a customized straw-like device (gustometer; **see Figure 1B**) attached to the head coil, allowing for precisely timed and measured delivery of stimuli^39^. Two versions of the milkshake task were created where predictive cues were counterbalanced as to what they predicted. Versions were consistent within participants and the same as their “mock scan.” Visual cues were projected on a screen behind the bore and viewed through a mirror for 2 seconds, followed by a jittered interstimulus interval (ISI) before 3mLs of milkshake or tasteless stimuli was delivered over 5 seconds; after a milkshake delivery a rinse of water was delivered ^24,29^. A variable jitter of 5-12 seconds followed stimulus delivery to allow for swallowing. A total of 18 stimulus trials for both milkshake and tasteless were completed for each run, and two runs occurred each session. Stimulus order, Inter Stimulus Interval (ISI), and Inter-Trial Interval (ITI) were determined using genetic algorithms for improved design efficiency^40^. PsychoPy (version 2023.1.3) was used to control all visual cues and pump triggers^41^.

## Statistical Analysis Demographics

Participant characteristics were summarized using the tableone package in R. Continuous variables were reported as mean (standard deviation), and categorical variables were presented as counts (percentages). Descriptive statistics were used to summarize participant characteristics, and comparisons (student’s t-test) were made between adolescent (18-21) and young adult (22-25) age groups. Statistical significance was defined as p < 0.05.

## fMRI

First, distortion correction and unwarping were performed using topup^42^ as implemented in FMRIB Software Library (FSL)^43^. Then, preprocessing was completed using SPM12 (Wellcome Trust Center for Neuroimaging, London, UK). Preprocessing consisted of realignment, co-registration, normalization, and smoothing (6mm full width half maximum gaussian kernel). First and second level analyses were completed using SPM12.

On the first level, box-car models for each stimulus were created (milkshake cue, tasteless cue, milkshake consumption, and tasteless consumption), and their temporal derivative. Rinse, framewise displacement (FD), DVARS, motion parameters^44^, and cerebrospinal fluid (CSF) signal fluctuation were entered as regressors of no interest. Contrasts of interest were created: milkshake cue > tasteless cue, milkshake consumption > tasteless consumption, and their inverses.

The first level analyses described here were then brought up to the group level using a within-subjects two-way ANOVA design with time (pre vs post) and diet (UPF vs non-UPF) as factors. To test our first hypothesis that brain response to milkshake would differ after the UPF diet condition, a two-way ANOVA with time (pre vs post) and diet (UPF vs non-UPF) was built and the diet-by-time interaction was tested. Previously, we tested all randomization factors for this trial (BMI, sex, and age) and reported an effect of age on intake^28^. Here to test these factors, we built a separate three-way ANOVA interaction model with time, diet, and each randomization factor: sex (male vs female), BMI (8.5-24.9 vs 25-29.9 kg/m), and age (18-21 vs 22-25) as factors. Post hoc comparisons were conducted on extracted parameter estimates using estimated marginal means (emmeans package in R) and Tukey’s Honest Significant Difference (HSD) method to control family-wise error rate.

To test the relationship between hypothesized intake outcomes, such as total energy (kcals) consumed at the *ad libitum* buffet, and brain response, we built separate two-way ANOVA diet-by-time interaction models described previously and entered each intake variable from the *ad libitum* buffet, EAH test, and participant’s habitual diet (**Table 2**), as a covariate. To further examine how diet-induced changes in brain response to milkshake are influenced by measures of intake, we subtracted brain response images from each session ([postUPF - preUPF] - [postnon-UPF – prenon-UPF]) to allow for regression analyses. Next, we subtracted each intake variable (**Table 2**) across conditions (UPF-non-UPF) to create a single value for each test, for each participant. These values were then regressed against the whole brain to examine if changes in brain response after diet condition correlated with any intake variable (**Table 2**).

As previously described, a single anatomical mask of *a priori* regions of interest (ROIs; striatum and prefrontal cortex) was applied using a small volume correction (pFWE < 0.05, see **Supplemental Figure 2**)^25,29,45^. Whole brain analyses were corrected at pFWE< 0.05. For improved visualization of the cluster containing the peak, all brain data are displayed with puncorr<0.001, k>10.

**Supplemental Figure 2:**
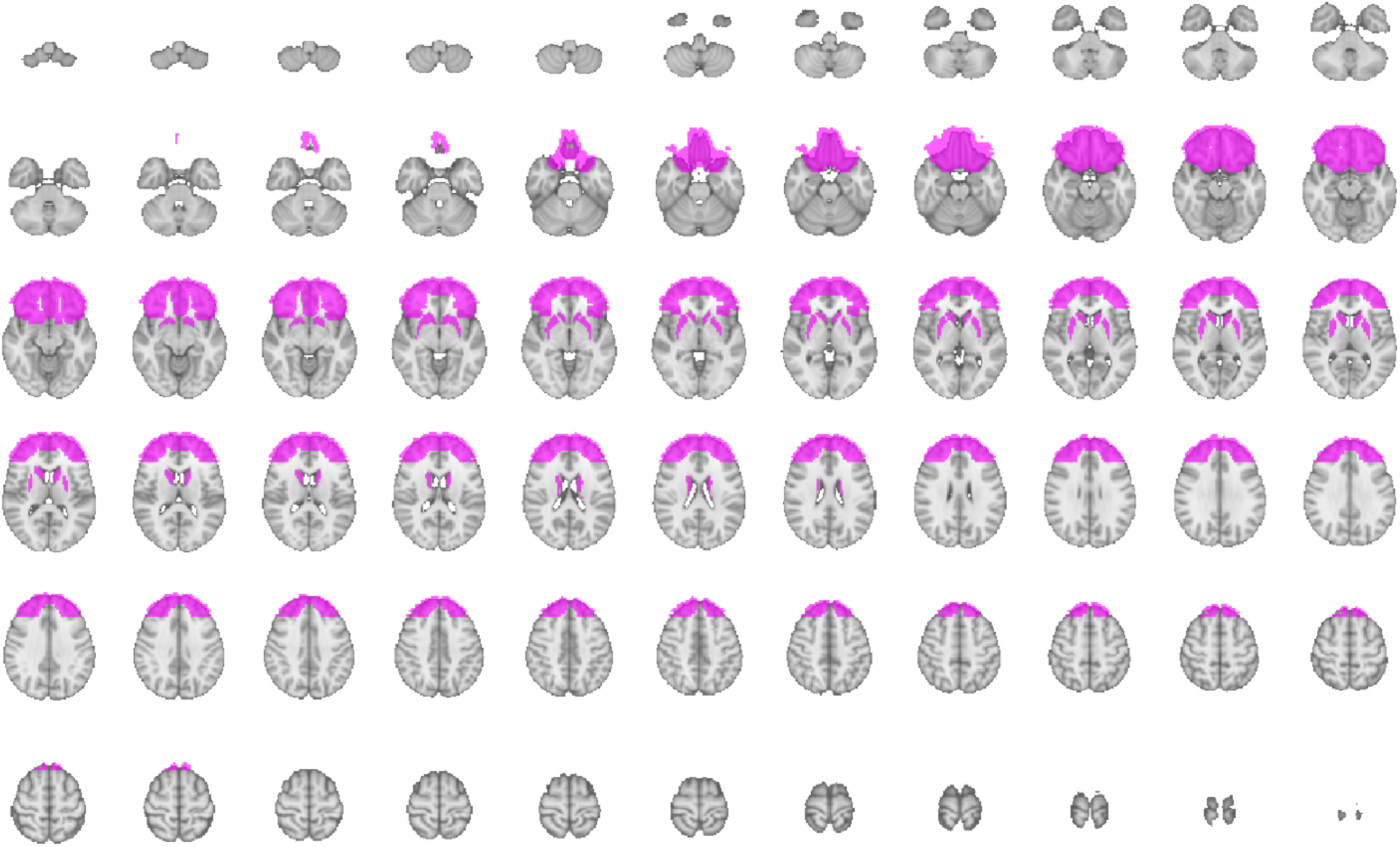
ROI mask. *A priori* region of interest (ROI) mask including the prefrontal cortex and striatum, depicted in axial mosaic.

## Results

### Participant Characteristics

The final sample presented here is n=22 (15 female; 16 Caucasian; **Supplemental Figure 1**). There were 7 participants (aged 18-21) in the adolescent age group, and 15 participants (aged 22-25) in the young adult age group. Participant characteristics are described in **Table 1**. Habitual diet as assessed from dietary recalls and energy intake at the *ad libitum* buffet meal and EAH test, reported previously^29^, is reported for the participants included in the fMRI analysis (**Table 2**).

**Table 1.** Participant Characteristics. BMI: body mass index, SD: standard deviation

|  | **Overall**<br>**(N=22)** | **Adolescent**<br>**18–21 (n=7)** | **Young Adult**<br>**21–25 (n=15)** | ***p***<br>**value** |
| --- | --- | --- | --- | --- |
| **Sex, n (%)** |  |  |  |  |
| Female | 15 (68.2) | 4 (57.1) | 11 (73.3) | 0.8 |
| Male | 7 (31.5) | 3 (42.9) | 4 (26.7) |  |
| **Age (years), mean (SD)** | 22.5 (2.0) | 20.3 (1.0) | 23.5 (1.3) | <0.001 |
| **Race, n (%)** |  |  |  |  |
| Asian/Pacific Islander | 4 (18.2) | 1 (14.3) | 3 (20.0) | 0.4 |
| White | 16 (72.7) | 5 (71.4) | 11 (73.3) |  |
| Other | 1 (4.5) | 1 (14.3) | 0 (0.0) |  |
| No answer | 1 (4.5) | 0 (0.0) | 1 (6.7) |  |
| **Ethnicity, n (%)** |  |  |  |  |
| Not Hispanic | 19 (86.4) | 6 (85.7) | 13 (86.7) | 1.0 |
| No answer | 3 (13.6) | 1 (14.3) | 2 (13.3) |  |
| **BMI (kg/m<sup>2</sup>), mean (SD)** | 23.6 (3.0) | 24.6 (3.1) | 23.1 (2.9) | 0.3 |
| **BMI category, n (%)** |  |  |  |  |
| 18.5–24.9 | 14 (63.6) | 4 (57.1) | 10 (66.7) | 1.0 |
| 25–29.9 | 8 (36.4) | 3 (42.9) | 5 (33.3) |  |
| **Weight (kg), mean (SD)** | 66.3 (10.3) | 69.9 (10.8) | 64.6 (10.0) | 0.3 |

**Table 2:** Habitual dietary intake and energy intake in the *ad libitum* and eating in the absence of hunger tests. SD: standard deviation, kcals: kilocalories, UPF: ultraprocessed food.

|  | Overall<br>(N=22) | Adolescent<br>18–21 (n=7) | Young Adult<br>21–25 (n=15) |
| --- | --- | --- | --- |
| **Habitual Dietary Intake, mean (SD)** |  |  |  |
| Energy intake, kcals/day | 1875 (440) | 2075 (498) | 1782 (393) |
| % UPF Grams | 24.7 (13.0) | 22.4 (18.5) | 25.7 (10.2) |
| ***Ad libitum* meal total energy intake (kcals), mean (SD)** |  |  |  |
| non-UPF diet | 639 (182) | 637 (240) | 640 (158) |
| UPF diet | 629 (196) | 712 (294) | 591 (125) |
| **Eating in the Absence of Hunger total energy intake (kcals), mean (SD)** |  |  |  |
| non-UPF diet | 159 (82) | 162 (88) | 157 (83) |
| UPF diet | 173 (100) | 224 (107) | 150 (91) |

**Supplemental Figure 1:**
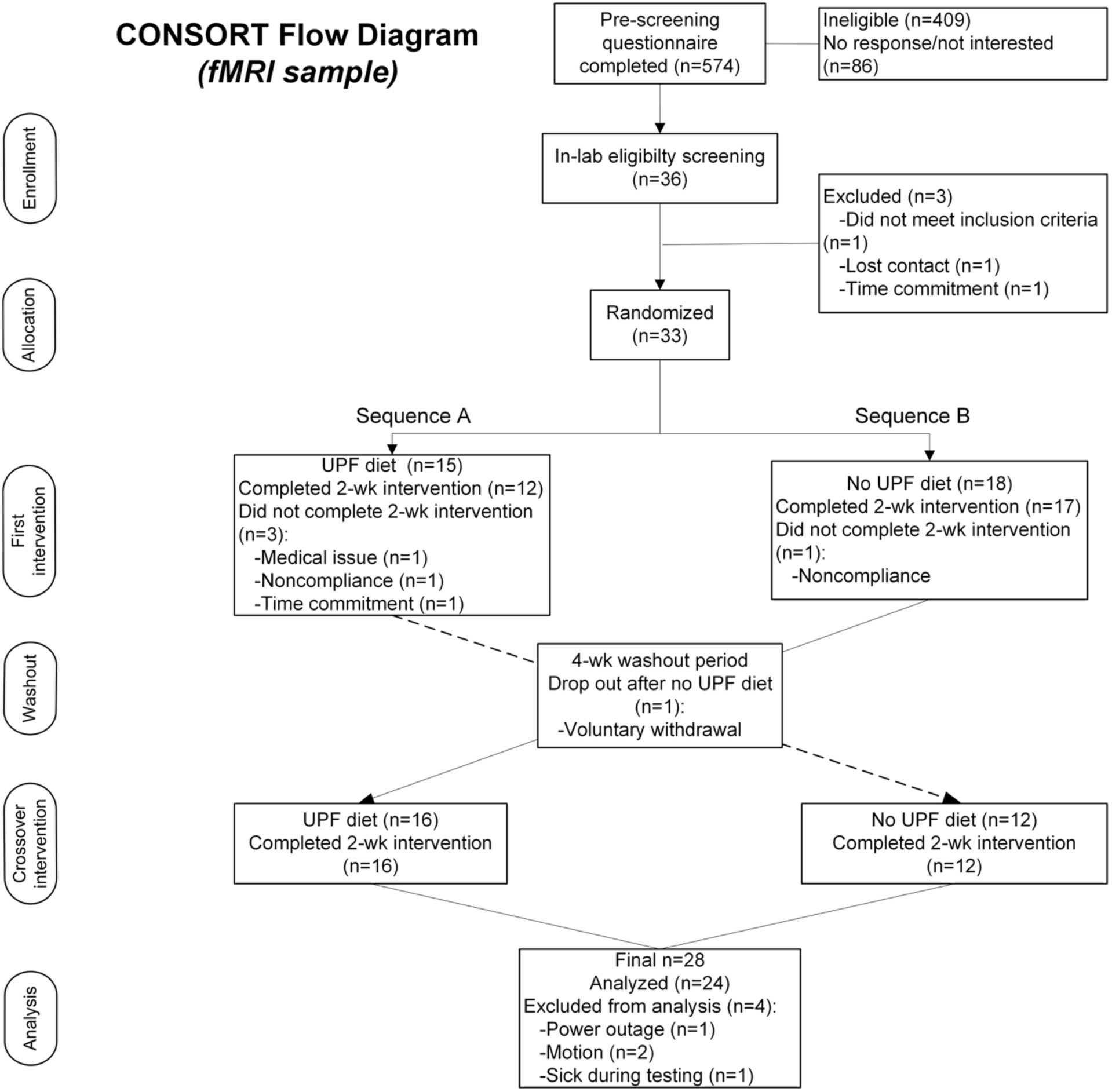
Consort Diagram showing participant flow from screening through completion and analysis for an ultraprocessed food trial (UPF).

### Intervention diet effects on brain response to milkshake cue and milkshake consumption

First, we examined if brain response to milkshake cue changed after each diet condition.

For this analysis, a diet-by-time interaction was tested using milkshake cue > tasteless cue contrast images from the first level analyses. We observed no significant voxels (p>0.05) for the diet-by-time interaction with milkshake cue (minus tasteless cue). These analyses were repeated for milkshake consumption, and again, no significant voxels survived.

### Intervention diet effects on brain response to milkshake cue and milkshake consumption by age sub-group

Next, we explored our randomization factors (age, sex, and BMI), as reported previously^28,29^. In our previous report, we observed significant effects of age but not BMI and sex on intake at the *ad libitum* buffet meal and EAH test. Here, we did not observe any significant voxels for a three-way interaction for milkshake cue (minus tasteless cue) with age, BMI, or sex. For milkshake consumption, again, no voxels survived multiple comparisons for an interaction with BMI or sex. However, we observed a three-way interaction with age in the orbitofrontal cortex (OFC), using our *a priori* ROI ([16, 28, -28], T=4.74, SVC p_FWE_ = 0.033; **Figure 2A**, **Table 3**). Visualization of parameter estimates (PE) revealed an interaction effect (t(60)=-3.18, p=0.002) where adolescents’ response to milkshake consumption, in the OFC, decreased after the UPF diet (t(60)=-2.91, p = 0.005) but increased after the NonUPF diet (t(60)=2.05, p = 0.045). Conversely, young adults showed no significant change across diets or timepoints, suggesting the effect of UPF consumption on brain response to milkshake varied as a function of age.

**Figure 2.**
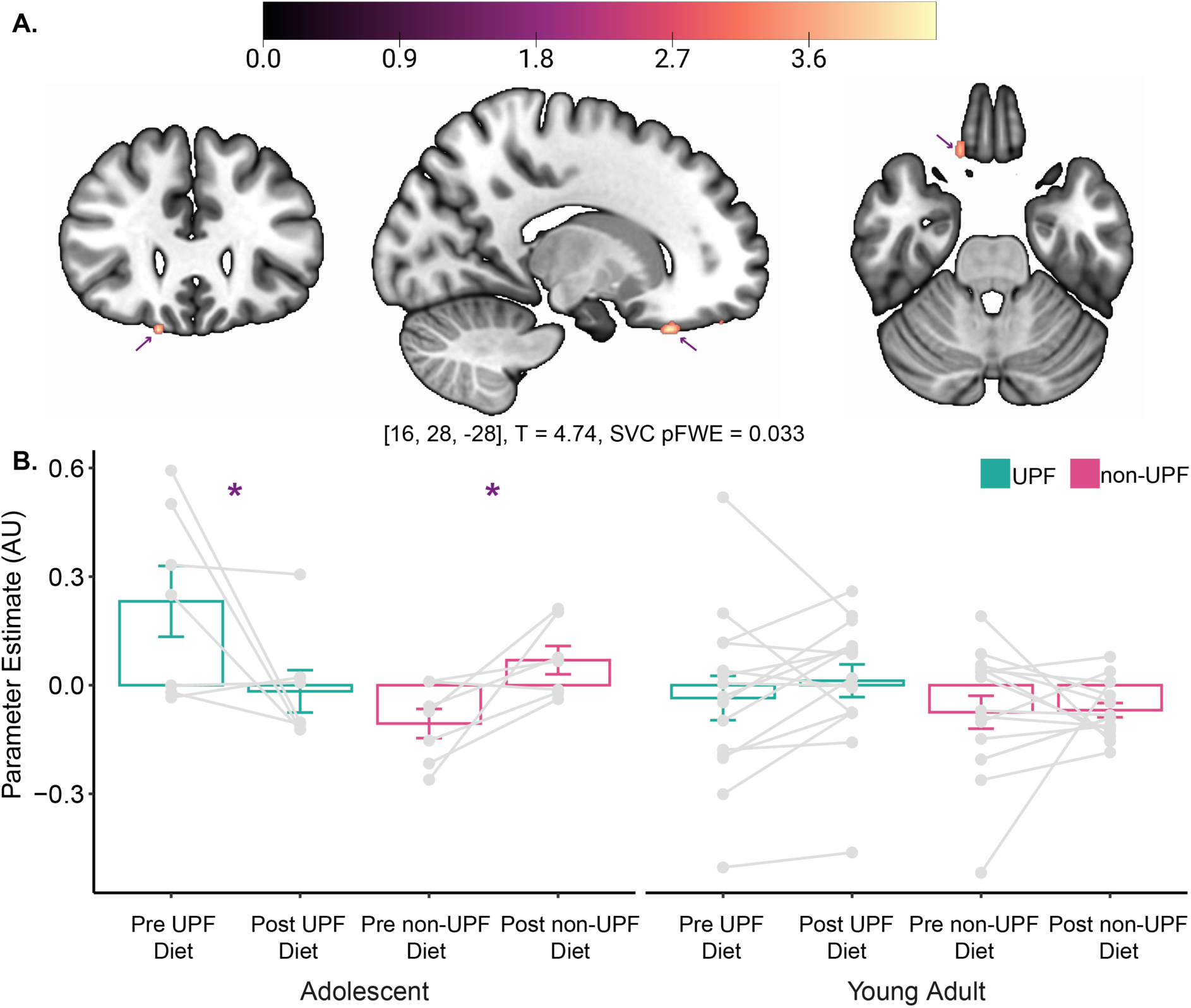
Three-Way Interaction: Diet, Time, and Age. **A.)** BOLD response in the OFC exhibited a Diet-Time-Age interaction elicited by milkshake consumption ([16, 28, -28], T=4.74, SVCp_FWE_ = 0.033). **B.)** Parameter Estimates were extracted from the OFC peak and plotted by diet (UPF vs non-UPF), time (pre vs post), and age (adolescent vs young adult). *p<0.05. SVC: small volume correction, FWE: family wise error correction, OFC: orbitofrontal cortex, UPF: ultraprocessed food, AU: arbitrary units

**Table 3.**
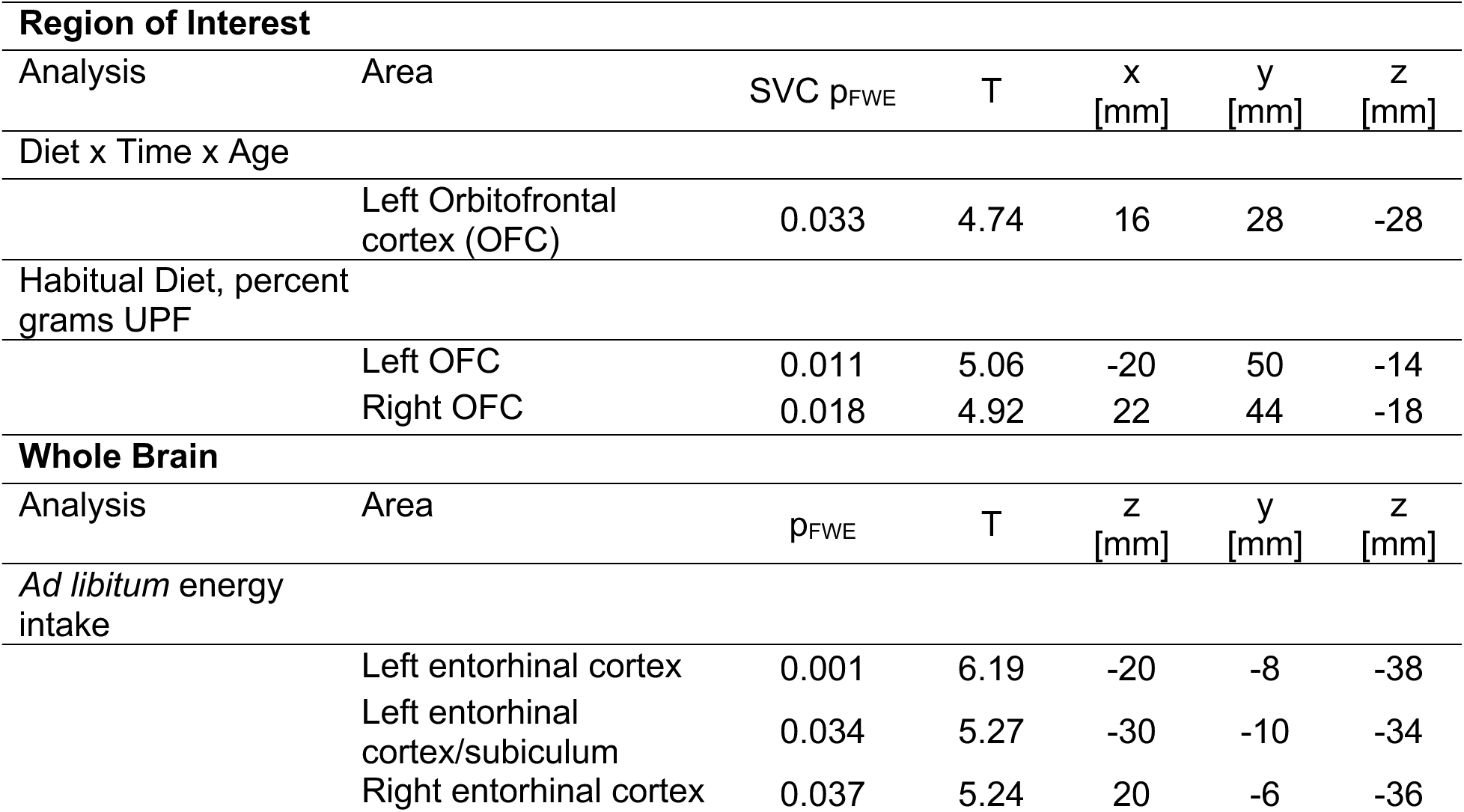
All fMRI results. SVC: small volume correction. FWE: family wise error correction. x, y, z: peak voxel location in Montreal Neurological Institute Coordinates.

| **Region of Interest** |  |  |  |  |  |  |
| --- | --- | --- | --- | --- | --- | --- |
| Analysis | Area | SVC $p_{FWE}$ | T | x<br>[mm] | y<br>[mm] | z<br>[mm] |
| Diet x Time x Age |  |  |  |  |  |  |
|  | Left Orbitofrontal cortex (OFC) | 0.033 | 4.74 | 16 | 28 | -28 |
| Habitual Diet, percent grams UPF |  |  |  |  |  |  |
|  | Left OFC | 0.011 | 5.06 | -20 | 50 | -14 |
|  | Right OFC | 0.018 | 4.92 | 22 | 44 | -18 |
| **Whole Brain** |  |  |  |  |  |  |
| Analysis | Area | $p_{FWE}$ | T | z<br>[mm] | y<br>[mm] | z<br>[mm] |
| *Ad libitum* energy intake |  |  |  |  |  |  |
|  | Left entorhinal cortex | 0.001 | 6.19 | -20 | -8 | -38 |
|  | Left entorhinal cortex/subiculum | 0.034 | 5.27 | -30 | -10 | -34 |
|  | Right entorhinal cortex | 0.037 | 5.24 | 20 | -6 | -36 |

To explore if OFC response changes linearly with age, we took the difference-in-difference contrast ([postUPF - preUPF] - [postnon-UPF – prenon-UPF]) and extracted the PE from the peak coordinates of the previous analysis. Linear regression of the PE against participant age revealed that there is a positive association between age and brain response to milkshake in the OFC *(R =* 0.68, *p <* 0.001), where OFC response to milkshake was higher as age increased, as shown in **Supplemental Figure 3**.

**Supplemental Figure 3:**
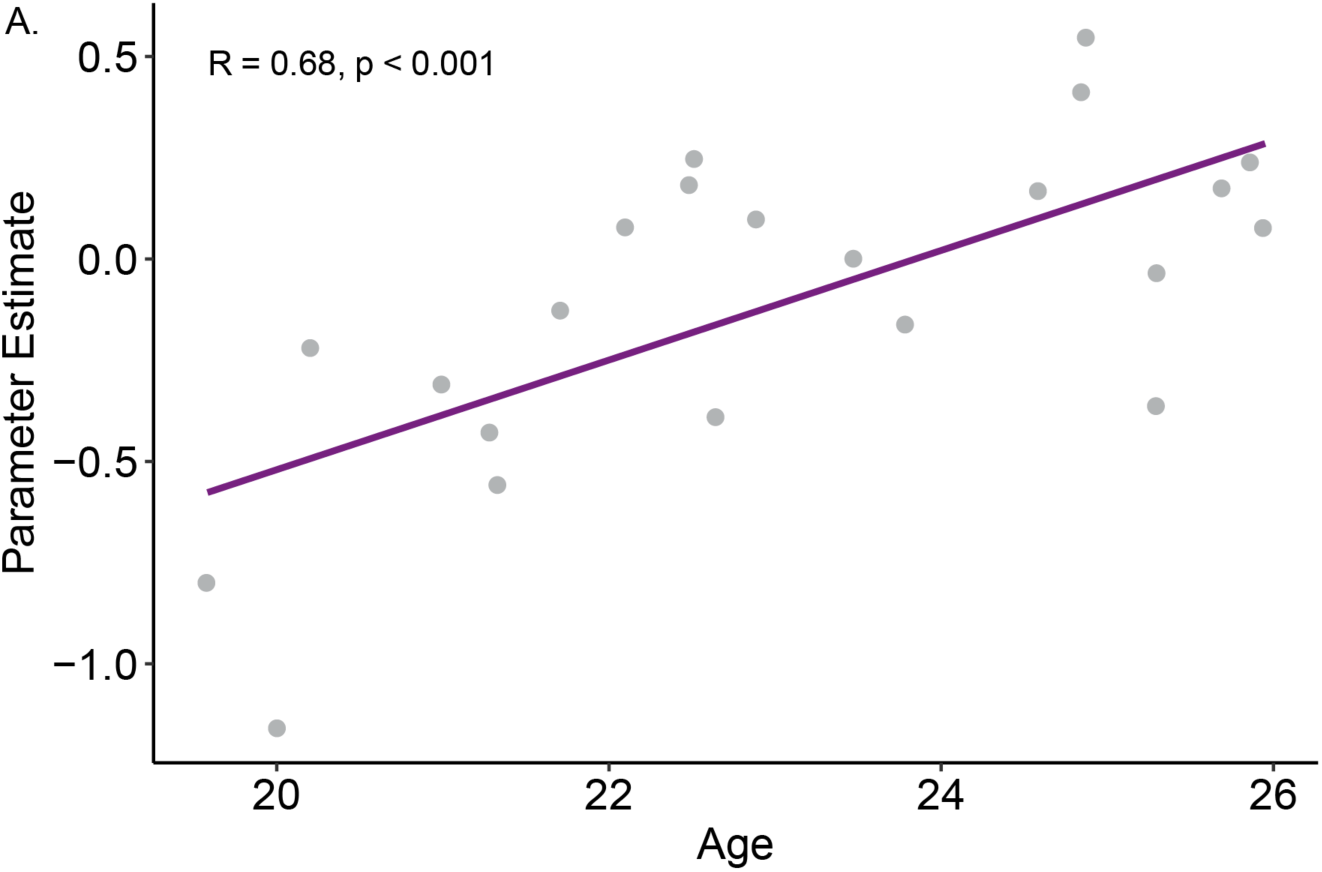
Orbitofrontal cortex parameter estimate positively correlates with age (R = 0.68, p < 0.001).

### Energy Intake in the Ad Libitum Buffet Meal is Associated with Brain Response to Milkshake

Recently, we published a report from this trial showing increased *ad libitum* energy intake after the UPF diet compared with the non-UPF diet in adolescents, but not young adults. We hypothesized that this change in behavior would be predicted by changes in brain response^29^. We found no correlation between energy consumed at the *ad libitum* buffet meal and brain response to milkshake cue. However, in response to milkshake consumption, we observed an inverse correlation between energy consumed at the buffet meal and brain response to milkshake in the bilateral entorhinal cortex ([-20 –8 –38], T=6.01, p_FWE_ = 0.002 and [20 –6 –36], T=5.24, p_FWE_ = 0.002; **Figure 3**, **Table 3, Supplemental Figure 4**), regardless of time or diet condition. Indicating that entorhinal response to milkshake could predict energy intake in a buffet meal regardless of recent diet history. Contrary to our hypothesis, we did not find that a difference in brain response to milkshake as a function of diet condition was associated with *ad libitum* energy intake; but rather, lower brain response to milkshake regardless of the previous diet condition preceded lower *ad libitum* energy intake at a later buffet meal.

**Figure 3.**
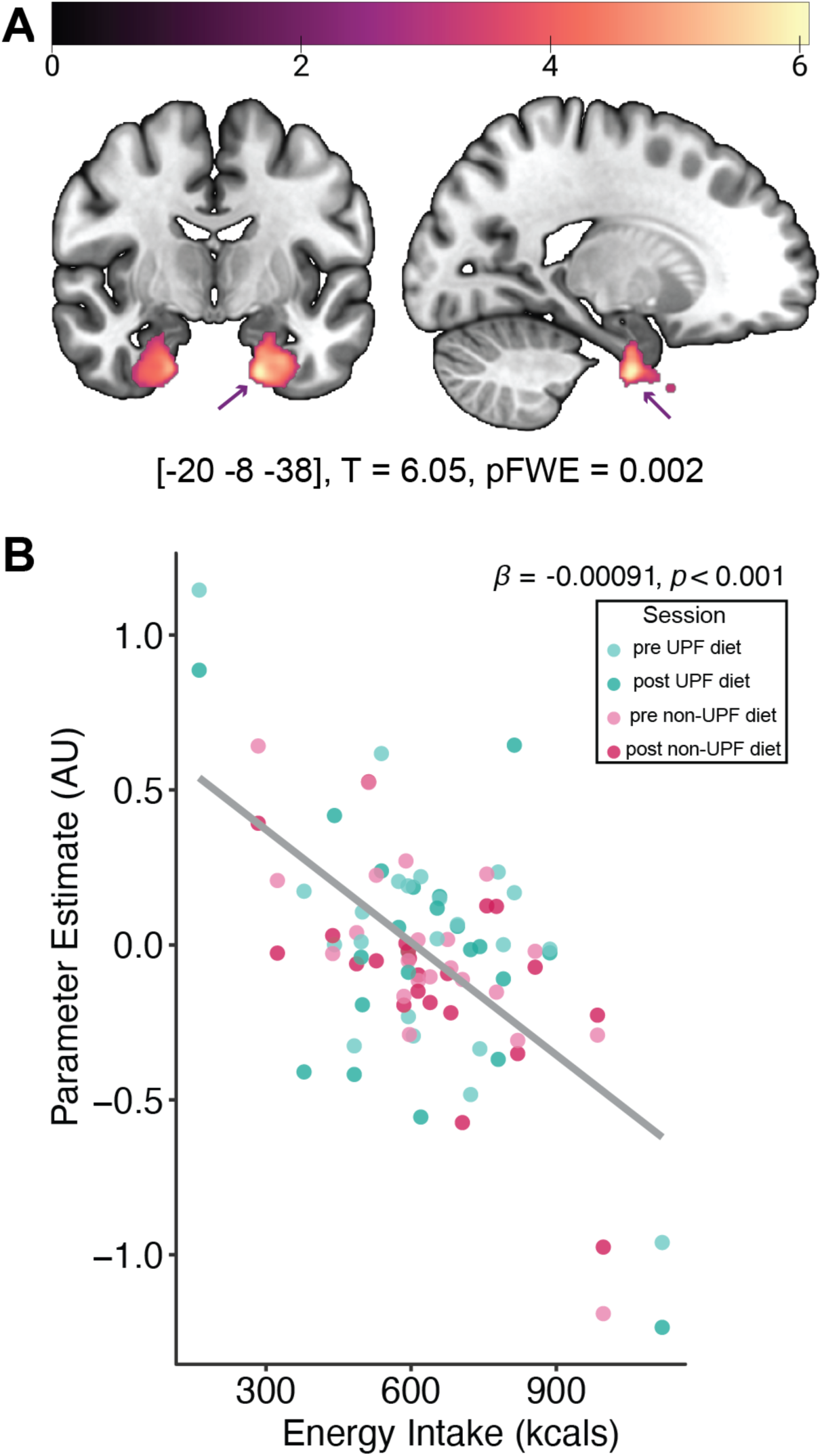
Ad libitum energy consumed is associated with brain response to milkshake consumption. **A.)** There was a negative correlation between kcals consumed at the *ad libitum* (ADL) buffet and brain response to milkshake consumption in the entorhinal cortex. **B.)** Visualization of the parameter estimates from the brain reveal lower activation in the entorhinal cortex is correlated with higher energy intake at the *ad libitum* buffet meal. UPF: ultraprocessed food, FWE: family wise error correction, AU: arbitrary units

Similarly, we tested UPF and non-UPF energy intake at the *ad libitum* buffet meal and UPF, non-UPF energy, and total energy at the EAH test and found no relationship with brain response to milkshake cue or consumption.

**Supplemental Figure 4:**
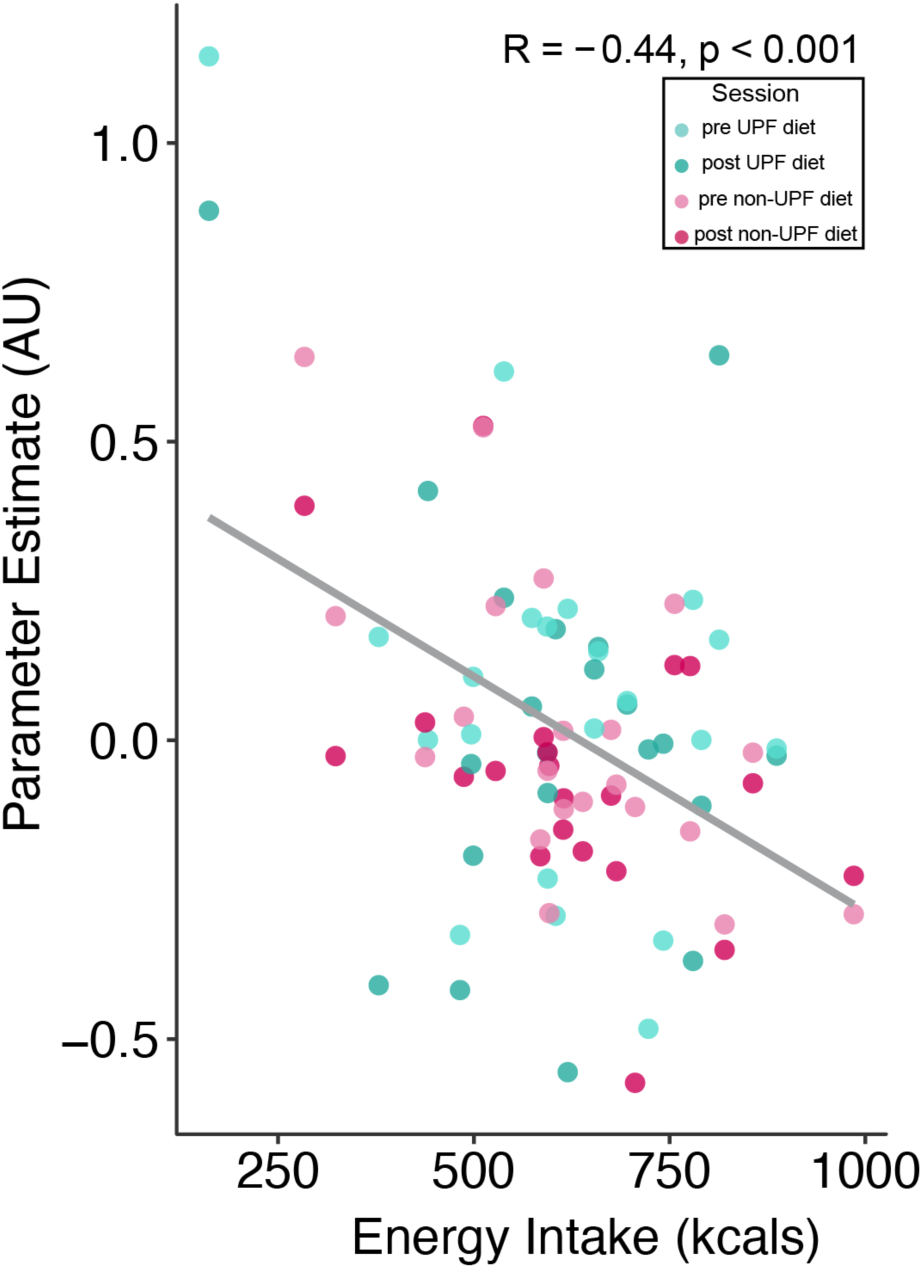
Entorhinal cortex activity correlates with intake at *ab libitum* buffet meal when excluding outlier (R = -0.44, p < 0.001).

### Habitual Diet is Associated with Brain Response to Milkshake

Although controlled feeding should mitigate the effect of a participant’s habitual diet, it is possible habitual diet, acting over the longer term, influences brain response even after controlled feeding. To test this, we entered each participant’s total UPF grams consumed derived from our dietary recalls as a covariate to the two-way ANOVA diet-by-time model described previously. We found no relationship between brain response to milkshake cue and habitual grams of UPF consumed. However, in response to milkshake consumption, we observed a positive correlation between habitual % grams UPF intake and brain response in the bilateral OFC ([-20 50 –14], T= 5.06, SVCp_FWE_ = 0.011, and [22 44 –18], T= 4.92, SVCp_FWE_ = 0.011 **Figure 4**, **Table 3**) where higher OFC response to milkshake was observed in participants with a greater proportion of habitual dietary intake from UPF. We do not observe an interaction with age and this effect (p>0.05). This indicates that habitual diet could modulate OFC response to milkshake over longer time scales, in all participants. However, some areas of the OFC are sensitive to acute intervention, when we entered habitual UPF consumption as an additional covariate in the three-way interaction of diet-by-time by age it did not change those results.

**Figure 4:**
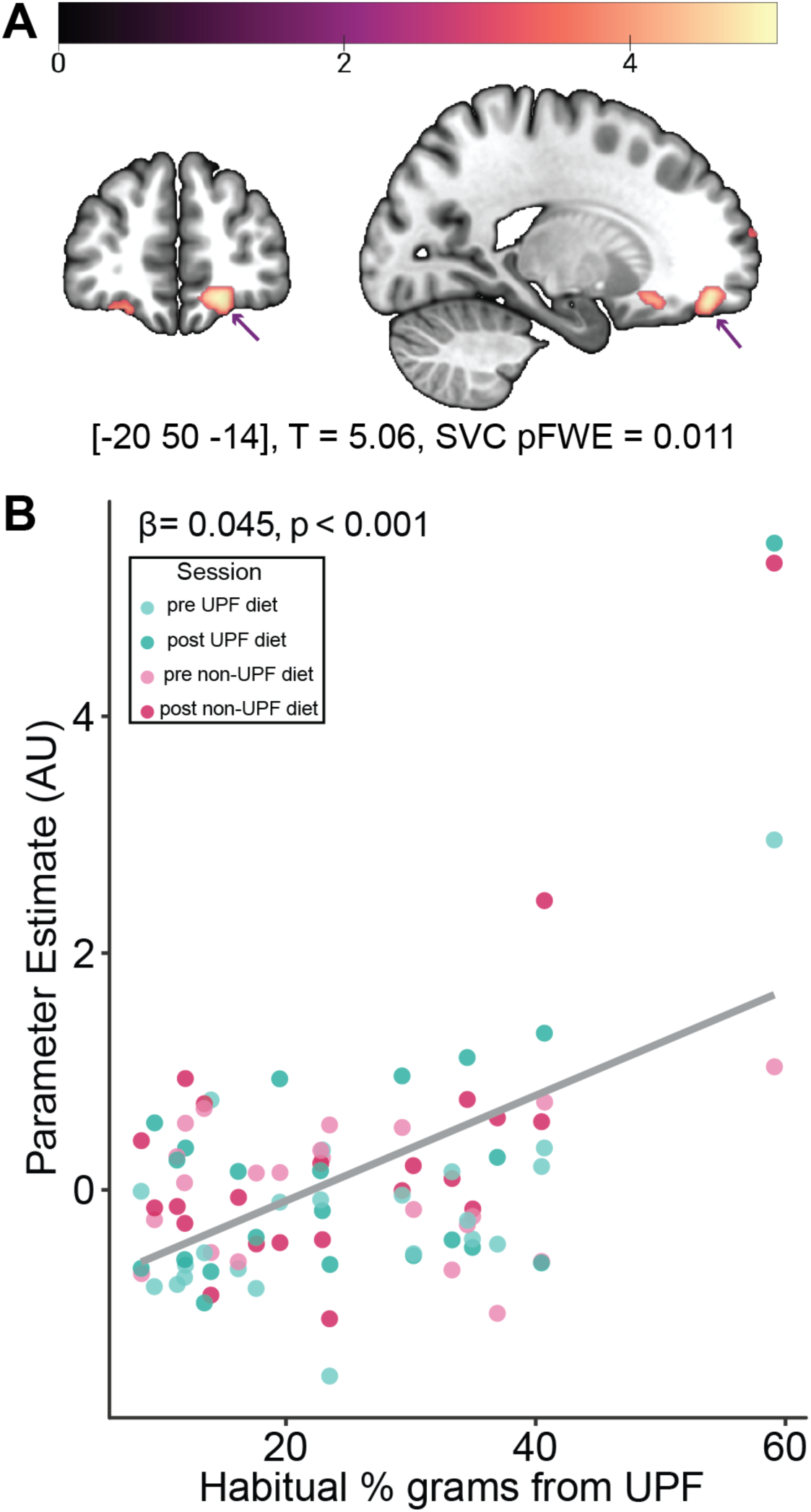
Percentage of UPF grams habitually consumed was positively correlated with orbitofrontal cortex (OFC) response to milkshake consumption: **A.)** There was a positive correlation between percent grams of ultraprocessed food (UPF) consumed habitually and brain response to milkshake consumption in the OFC. **B.)** Visualization of parameter estimates from the brain reveal higher percent of UPF grams habitually consumed was positively correlated with OFC brain response. SVC: small volume correction, FWE: family wise error correction, OFC: orbitofrontal cortex, UPF: ultraprocessed food, AU: arbitrary units

## Discussion

This trial investigated the effects of a high-UPF diet compared with an eucaloric, nutrient-matched diet containing zero UPF on brain response to a rewarding milkshake stimulus. We observed that overall, there were no effects of diet intervention on brain response to milkshake. However, when we examined our randomization factors, we found that OFC response to milkshake consumption was reduced after a UPF diet but increased after a non-UPF diet in the adolescent age group but not the young adult group. This pattern of results is similar to previously reports from this trial, where adolescents ate more in *ad libitum* test meals after the UPF diet than after the non-UPF diet, but not young adults^28^. We did not observe a correlation between change in brain response over the diet periods and intake, as hypothesized. But rather, regardless of age or diet condition, entorhinal response to milkshake consumption was inversely correlated with the total energy intake at the *ad libitum* buffet meal in that lower entorhinal cortex activity preceded higher energy intake at the buffet meal. Finally, given no overall effects observed in our short-term intervention, we explored whether longer-term UPF intake before the controlled diet intervention influenced brain response and found it was positively correlated with brain response in the OFC, independent of diet condition or age.

Our observed effects in the OFC were influenced by habitual diet, acute dietary change, and age. The OFC is a hub for food valuation, contains the secondary taste and olfactory cortex, and is still under development into the third decade of life^26,46–49^. Evidence from primates and humans demonstrates that the OFC is the integration point where visual, olfactory, gustatory, and somatosensory pathways converge to encode reward value and pleasantness of foods^46–50^. Interestingly, responses in the OFC exhibit sensory-specific satiety, where a selective decrease occurs to a food that was just consumed, but remains responsive to different foods^46,51^. It is possible that sensory specific satiety occurred more rapidly in the adolescent age group after the UPF diet in response to a UPF milkshake. OFC function could be more sensitive to perturbations induced by dietary change in the younger group given this region is undergoing rapid developmental change^26,47^. Interestingly, we observed acute changes in OFC function in response to diet change and differences in OFC response to milkshake consumption that correlated with habitual proportion of dietary UPF. The latter is consistent with dietary manipulation effects on OFC dynamics in rodent models^52^, where both acute and long term effects are observed. It is possible while our younger participants were perhaps more sensitive to the acute intervention, longer term dietary patterns still influence OFC function in all ages tested. These correlations were observed in different subregions of the OFC which are hypothesized to have different functions^53,54^. Potential mechanisms for the observed changes beyond nutrient composition include additives and advanced glycation end-products (AGE), which have been shown to accumulate in the brain^50,55–60^.

Contrary to our hypothesis, we did not observe changes in brain activity as a result of dietary intervention predicted eating behavior, but rather activity in the entorhinal cortex correlated with energy intake at a subsequent buffet meal regardless of the diet condition. Brain response to foods and food cues in the nucleus accumbens^61^, hypothalamus^56^, OFC^56,62^, amygdala^62^, and the midbrain^63,64^ has been previously demonstrated to be predictive of later food intake. The entorhinal cortex receives projections from many of the areas previously shown to predict food intake and its major output structure, the hippocampus, is thought to guide eating behavior by integrating past food experiences, internal states, and contextual cues^65^. These findings expand the functional network of brain areas that predict later intake.

To date, no trials have investigated the influence of UPF exposure on brain response to UPF in adolescents and young adults. This research addressed this gap using a rigorously designed crossover controlled feeding approach with well-characterized, nutrient-matched diets that differed only in level of food processing^28,29,66^. Dietary quality, total and non-beverage energy density were matched between the two diets. *Ad libitum* buffet meal food items consisted of UPF and non-UPF foods and beverages matched by food group type (e.g., 100% orange juice vs Sunny Delight juice drink; homemade banana muffin vs Pop-Tart) and similar in texture and energy density^29^. In addition, all observed changes in brain response occurred despite body weight stability^28^. However, this trial was powered to observe effects in the overall groups, not in the randomization factors tested here and previously^28^. Therefore, these findings should be repeated in a larger sample. It will also be important to understand if these brain effects persist when participants return to their regular diet, rather than a cross-over controlled diet. Given the ongoing maturation of the prefrontal cortex, including the OFC, during emerging adulthood, it will be particularly important for future studies to empirically test whether UPF consumption impacts executive functioning. Finally, it will be important to examine these potential effects in younger participants and utilize longer controlled feeding periods.

In conclusion, these findings provide evidence that food processing independent of nutrient content may only acutely influence neural response to foods during adolescence. Importantly, these same brain regions are still undergoing maturation and may represent a window in which the brain is particularly sensitive to UPF exposure.

## CRediT Statement

EHL: Data curation, formal analysis, investigation, methodology, resources, software, visualization, writing—original draft; MLR: Data curation, investigation, methodology, resources, writing—review & editing; MA: methodology, data curation, formal analysis, validation, writing—review & editing; WY: Data curation, investigation, resources, writing—review & editing; MEB: data curation, validation, writing—review & editing; AG: data curation, validation, writing—review & editing; RS: data curation, validation, writing—review & editing; HL: data curation, validation, writing—review & editing; RK: data curation, validation, writing—review & editing; DLH: data curation, validation, writing—review & editing; VEH: conceptualization, writing—review & editing; KPD: conceptualization, writing—review & editing; BK: conceptualization, writing—review & editing; BMD: Conceptualization, Funding acquisition, methodology, project administration, supervision, writing—review & editing; AGD: Conceptualization, Funding acquisition, methodology, project administration, software, supervision, writing—review & editing

## Data Availability

All data and code will be made publicly available upon peer-reviewed publication of this manuscript.

## Acknowledgements

This study was funded by R21HD109722 to AGD and BDM. We would like to thank all of our participants for their contribution to this study.

